# Estimation of true number of COVID-19 infected people in Japan using LINE questionnaire

**DOI:** 10.1101/2020.04.15.20066100

**Authors:** Shin-ichoro Tanaka, Shinya Oku

**Affiliations:** The Institute of Scientific and Industrial Research, Osaka University, Ibaraki, Osaka 567-0047, Japan

## Abstract

The authors estimated the true number of COVID-19 infected people in Japan using the LINE questionnaire data and the PCR test results. A statistically significant correlation was observed between the infection rate per prefecture with PCR test and the rate of high fever. Using this correlation, true number of COVID-19 infected people in Japan was estimated approximately twenty thousand (±ten thousand) as of April 1, 2020.

## I. INTRODUCTION

COVID-19 has been evolving worldwide in the first half of 2020 and also in Japan, the confirmed number of infected people was reported as 5,438 on the website of Ministry of Health^1^, Labour and Welfare (MHLW) as of 11 a. m. of April 12, 2020. This number, confirmed by Polymerase Chain Reaction (PCR) tests, only shows the subpopulation of the infected and its true number is presumably more than this. While the estimation of true number is very important in order to prevent from so-called “collapsed healthcare system “or overwhelming hospitals as well as in order to acquire herd immunity in the future, it is challenging to estimate the true number with the number of tested positive. Its difficulty comes from 1) the extremely limited number of PCR tests with various reasons and 2) as PCR test is performed when the subject is suspicious of having COVID-19 infection with certain medical situation such as high fever or continuous general fatigue, etc., sampling of PCR test is not randomized. That being understood, if the PCR test subjects are randomized and the number of PCR tests is sufficient, by calculating the product of national population and (confirmed number of infected)/(number of PCR tests), a satisfactorily reliable true number of infected can be obtained. However, since only the people suspicious of being infected undergo the tests actually the positive rate is much higher than that of the randomized samples. Using the current (confirmed number of infected)/(number of PCR tests) leads to estimate the true number quite excessively. For example, as of April 9, 2020, MHLW website^2^ states that 4,667 out of 54,284 was tested positive. If this rate is multiplied by 126 million of entire Japan population, approximately 10 million is the assumed true number of the infected, which would not reflect the actual situation. Under the status quo as such, a large-scale randomized questionnaire (1st) by MHLW/LINE corporation was conducted from March 31 to April 1, and the result was made open^3^ on April 10. The authors analyzed these data statistically and estimated the true number of COVID-19 infected patients.

## II. MATERIALS AND METHODS

As the fundamental data, the following three were used.

1. Questionnaire data by MHLW/LINE performed during March 31 and April 1^3^ Mainly was used the number of high fever people per prefecture and the number of responders to questionnaire.
2. Data from J.A.G JAPAN cooperation^4^ Using the file^5^ available at https://gis.jag-japan.com/covid19jp/, the confirmed number of PCR positivity per prefecture was calculated and the latent periods of the infected were estimated using the dates of initial symptom(s) and the confirmation date of PCR positivity.
3. Population per prefecture by National Population Census 2015.^6^

With these datasets, the following steps were performed. Figures will be described in detail afterwards.

- Fig. 1 showed the ratio of patients with high fever over the questionnaire responders per prefecture using the above 1)

**FIG 1.**
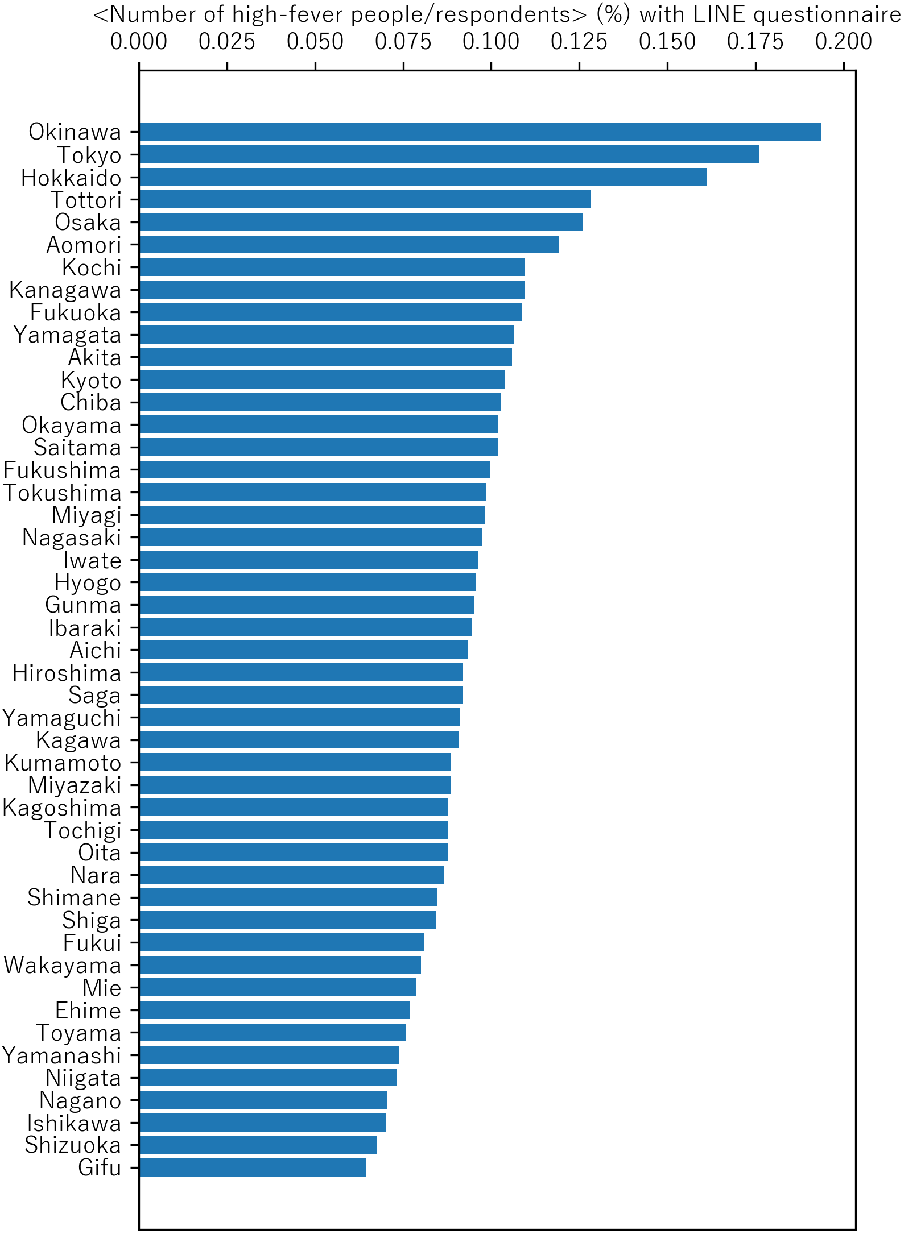
⟨Number of high-fever people/respondents ⟩ (%) per prefecture with LINE questionnaire.
- Fig. 2 showed the distribution of dates from initial symptom to infection confirmation using the above 2)

**FIG 2.**
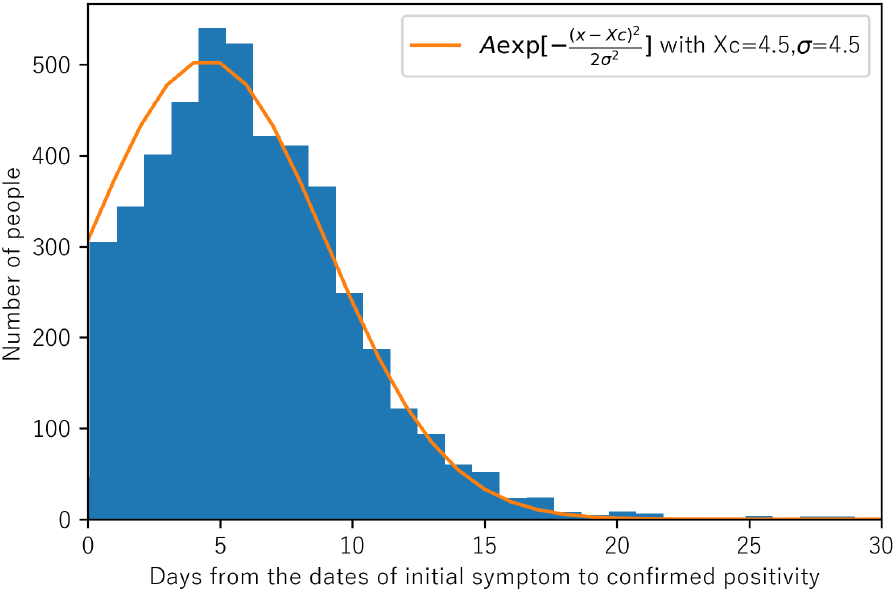
Distribution of days from the dates of initial symptom to confirmed positivity.
- Fig. 3 showed the ratio of accumulated number of PCR positivity over population per prefecture as of April 5 (this date was deducted from Fig. 2.)

**FIG 3.**
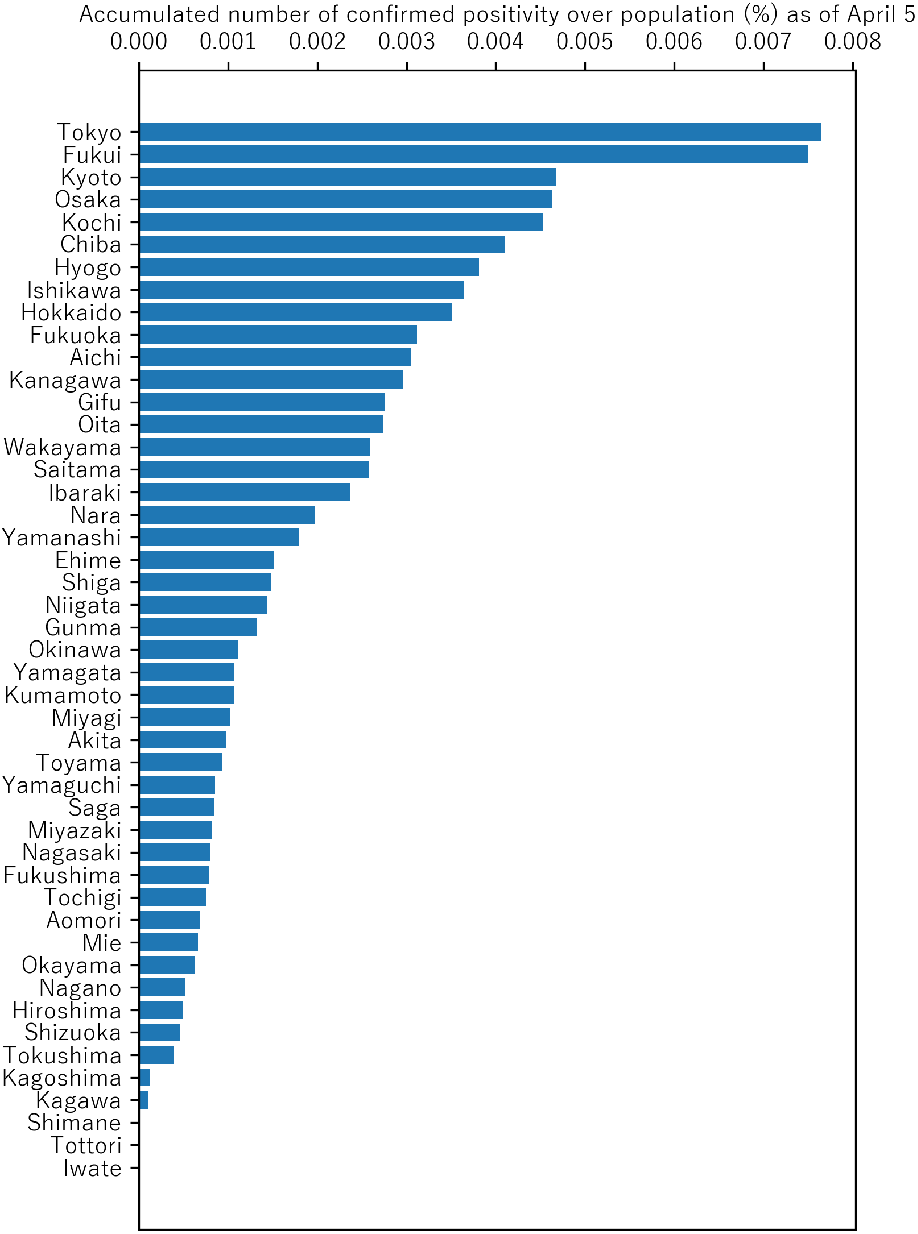
Accumulated number of confirmed PCR positivity over population (%) per prefecture as of April 5.
- With Fig. 4, on the basis of Fig. 1 and 3., (a) determined the correlation between the ratio of accumulated number of PCR positivity over population and the ratio of patients with high fever over the questionnaire responders per prefecture, (b) performed linear regression analysis, (c) identified the component of COVID-19 over entire high-fever subjects and then (d) estimated the true number of COVID-19 patients nationwide.

**FIG 4.**
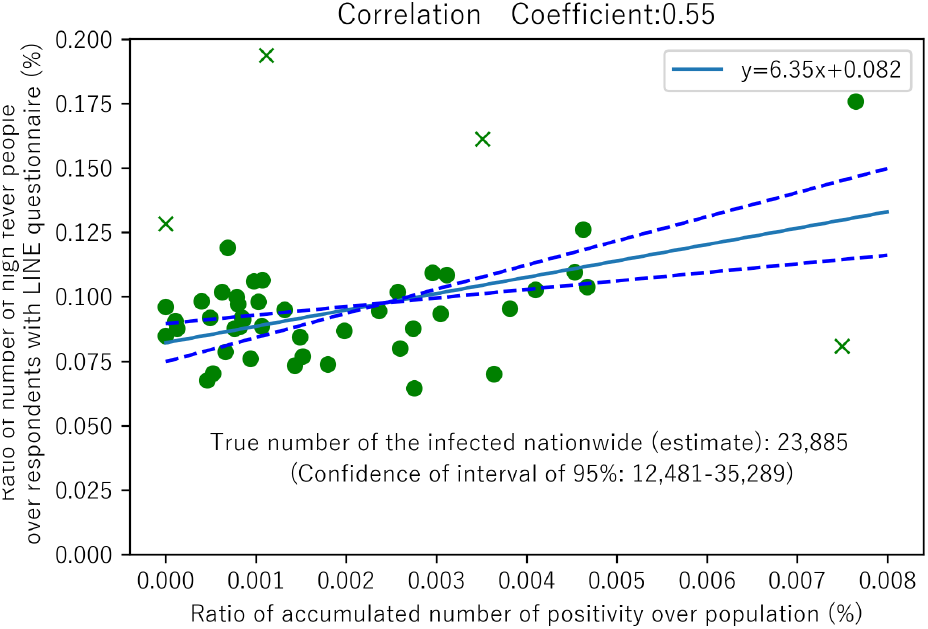
Correlation between ⟨accumulated number of SARS-CoV-2 positivity (as of April 5) over population⟩ and ⟨number of high fever people / respondents with LINE questionnaire⟩.
- With Fig. 5, taking the influence of recovered patients into account, identical analyses with Fig. 4 were performed.

**FIG 5.**
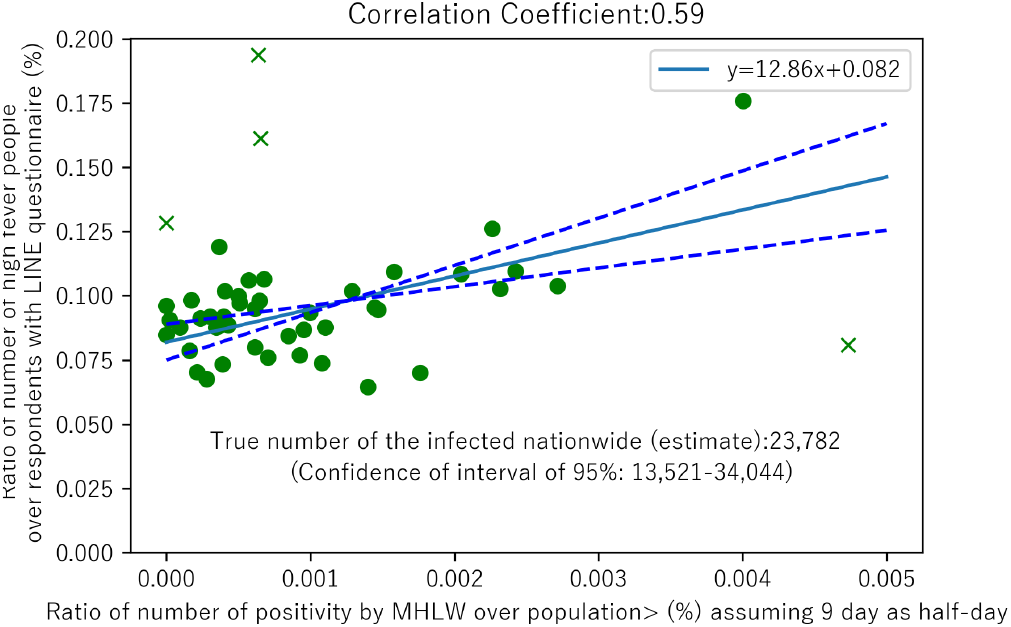
Correlation between ⟨number of SARS-CoV-2 positivity (as of April 5, assuming 9 day as half-day over population) ⟩(%)⟨number of high fever people over respondents with LINE questionnaire⟩.

Anaconda Python 3.7^7^ was employed to acquire data from the Internet, analyze and draw graphics. Regarding the regression analysis, the software library called statsmodels^8^ was employed and Ordinal Least Squares (OLS) algorithm was applied.

## III. FUNDAMENTAL DATA AND RELATED METHODOLOGY

Firstly, LINE questionnaire will be described here. As discussed in the report by MHLW, high fever caught in the questionnaire is considered as including the fever with COVID-19 as its cause. In contrast with the ongoing PCR test, this questionnaire is quasi-completely randomized,^9^ exhibiting its large scale of 23,992,701 replies. This means that these data can be served as the fundamental data to deduce the true number of infected. As it should be noted that the data contains the fever with other reasons than COVID-19, the data itself could not be regarded as the number of the infected. The authors underpinned the rate of high fever per prefecture, which were served as the data source for estimating true number of the infected.

Figure 1 exhibits the number of high fever/responses per prefecture by LINE questionnaire. Bar charts were provided after sorting data in descending order. Except for the three prefectures which showed the extremely high numbers, the difference among the rest remains within a certain range. As reported in the news, the spread of COVID-19 infection is not homogenous nationwide; i. e., in Tokyo, only seldom infections were found when many were found in Hokkaido in its initial days. Therefore, it was expected that the distribution of high-fever patient caused by COVID-19 shows a similar distribution to the confirmed cases according to the regions visibly. In contrast, the distribution of high-fever patient caused by non-COVID-19 reasons was considered as stable regardless of the regions; that the identical distribution, with some extent of statistical error, could be expected. If the variations according to the regions could be taken into account, the part of the high-fever patients caused by COVID-19 could be extracted. In order to evaluate the appropriateness of this model, the comparison between the national distribution of PCR positive patients at the timing of LINE questionnaire and that of high-fever patients assessed in LINE questionnaire, would be effective. It could not be expected to determine the true absolute patient number with PCR test data, however, the relative distribution reflects the true patient distribution properly, assuming that the philosophy to design PCR test per prefecture remains identical nationwide. With this consideration, the authors could believe that the simple comparison between the distributions of PCR positivity and LINE high-fever suffice. Needless to state, when pursuing the trend of positive patients, not only the regional difference but also the time-dependent change of a patient history from the infection via the latent period to the appearance of initial symptom, should be considered. Simply applying the current statistical data as the data for infected might cause a loss of accuracy; rather the data at the exact timing of LINE questionnaire should be applied. Furthermore, as the fixation of number of the infected happens certain days later from the infection, waiting for the continuous symptom(s) to decide to perform PCR test, not the confirmed number of a date, but the newly symptomatic patient number of the same date should be used. Meanwhile, as the information of confirmed number of patients at a certain date includes only the limited number of initial symptom dates, the exclusive use of these data makes the analysis statistically non-robust.

With this consideration above, the authors conducted an additional analysis using the data investigated and made open by J.A.G. Japan^5^ displayed in Fig. 2. This dataset has the data of initial symptom and those of confirmed positivity (as of April 12). As the date of initial symptom was not identified for every patient, this data showed only the identified data, meanwhile, satisfactorily statistical distribution was exhibited. The authors superimposed the Gaussian distribution onto this data (where × is the difference between dates of initial symptom and confirmed positivity). Although this distribution model was not completely concordant with the data and there might exist some other better distribution which fits more, it was shown that there are 4 to 5 days of difference between the dates of initial symptom and confirmed positivity. It could be reasonable to use the confirmed positivity data on April 5 to be compared with LINE questionnaire data on March 31 to April 1.

Fig. 3 displayed the accumulated number of confirmed positivity on April 5 per prefecture, divided by the population obtained from the National Survey 2015. It shows the fact that the rates of COVID-19 positivity varies largely with each prefecture. As the next step, the authors investigated the correlation between the distribution of positivity rate per prefecture and that of high-fever rate.

## IV. DATA ANALYSIS AND ESTIMATED TRUE NUMBER OF THE INFECTED

Fig. 4. is the scatter graphic setting the accumulated number of confirmed positivity divided by population on April 5 as horizontal axis and ratio of number of people declaring high-fever over the number of total responders as vertical axis. The data from 4 prefectures shown as × in the figure, i.e. Tottori/Okinawa/Hokkaido/Fukui, from left to right) were apart from those of the other 43 prefectures. Putting Hokkaido aside, the 3 prefectures have smaller populations and incidental increase of infection and/or high-fever might arise. It was not quite explicable that Hokkaido is out of the general order, however, its vast surface area and extremely low population density other than Sapporo and some other outstanding cities might influence and its comparison with other prefectures remains challenging. At any rate, as excluding the extraordinary observation data is a common statistical technique, the authors decided to use the remaining 43 prefectures. As a further note, Tokyo, which has a combination of high accumulated number of the positivity and high rate of high-fever, with the thoughts that the population is large enough not to be excluded, the authors dare not exclude and continue to use this despite its plot on the graphic.

Pearson’s correlation coefficient *r* between ⟨ratios of accumulated positivity over population for 43 prefectures⟩ (%) and ⟨ratios of high-fever people over questionnaire responders for 43 prefectures⟩ (%) was calculated as *r* = 0.55. This means that the rate of PCR positivity and high-fever rate by LINE had an explicit correlation, showing the linearity with the slope of 6.35 and the intercept of 0.082 calculated by the least square method. Furthermore, the analysis with statsmodels library^8^ proved that the slope values range 3.32-9.98 and the intercept values 0.074-0.090 with the significance level of 0.05. The lines with these values were displayed by dotted blue lines in Fig. 5. As a note, if all the 47 prefectures were taken into the calculation, the correlation coefficient was as low as 0.28. With this value, while the use of data from all 47 prefectures seems inappropriate for the purpose of this paper, the authors could still confirm the relationship between these two ratios.

These results could enable to calculate the true number of the infected. With any given prefecture *p*, number of high fever (*N*_*fp*_) consists of the high-fever by COVID-19 infection (*N*_*Cp*_) and that indifferent to COVID-19(*N*_*fnp*_). Note that *N*_*fp*_ = *N*_*fnp*_ + *N*_*Cp*_. The quotient of *N*_*fp*_ by the population of prefecture *p* (*P*_*p*_), i. e., *N*_*fp*_/*P*_*p*_, is the rate of high-fever. Taking that LINE questionnaire sampling is randomized and large enough, into account,^9^ it could be safely concluded that the quotient *N*_*fLp*_/*N*_*Lp*_, i.e., the number of high-fever people with LINE questionnaire (*N*_*fLp*_) divided by the responses (*N*_*Lp*_), is equal to *N*_*fp*_/*P*_*p*_. Therefore, the following equation was deduced.

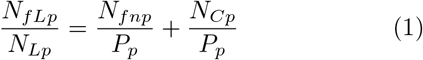

Herein, with the above linearity, putting *a* and *b* as the slope and intercept of regression line respectively and *N*_*T*_ *p* as the confirmed number of the infected, the relationship (2) was shown.

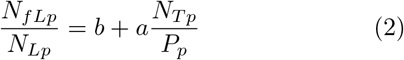

As (1) and (2) is true for any prefecture 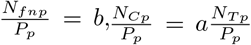, are true, hence the following relationship (3) is obtained.

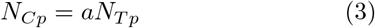

By summing up for all the 47 prefectures for both sides of equation, (n. b.: 4 prefectures excluded to calculate the coefficient were reverted, as they are not influential over the overall result.), we obtain

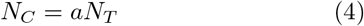

Here, *N*_*C*_ is the number of people with high-fever by COVID-19 as of April 1 and *N*_*T*_ is the confirmed PCR positivity number. With the use of “*a* “value in Fig. 4 and number of PCR positivity at April 5 (3,764 patitents^5^), estimation of 23,885 people was obtained as the true number of the infected who had high-fever nationwide on April 1, the very day of the LINE questionnaire (1st). Also, the confidence interval proven for “*a* “, the true positivity number was proved to be between 12,481 and 35,289 with the same significance level of 0.05.

In the analysis stated above, the authors used the accumulated number of confirmed positivity as of April 5 as PCR positivity. As it must be recognized that any patient infected would recover (or die) with time, simple sum of the infected is considered as larger than the infected at the time point. However, as the methodology to handle mathematically the decrease of the once infected patient by recovery or death is still under investigation, the authors decided to use the following method alternatively. Same as SIR model^10^, applying half-life of COVID-19 as *τ*_*HL*_ and letting *N*_*T P*_ as confirmed number of positivity, the following equation was assumed:

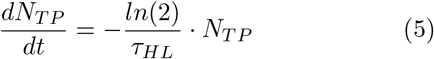

Here, setting the number of new PCR positive patients on day “*i*” of observation as *N*_*Ii*_, on day “*n*”, the number becomes 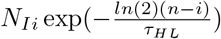), hence the actual number of confirmed positivity *N*_*T P*_ is as follows.

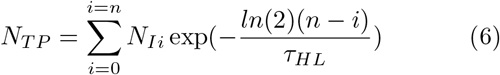

Using this equation and calculating the original data5 to produce the confirmed positivity as of April,^5^ their correlation was plotted as Fig. 5.

Here, the half-life is set as 9 day, which brought slightly improving regression coefficient, from 0.55 to 0.59 showing that the hypothesis applied here are reasonable to a large extent. As the change of patient number from” accumulated “to “non-accumulated” downwards, the estimation for the true number of the infected, adding consideration of half-life, was 23,783, producing no outstanding difference from the number of the precedent method. Despite this observation, with the increase of correlation coefficient and decrease of statistical error, the new range was 13,522-34,044 with the same significance level of 0.05, resulting in a slight improvement of precision. To confirm the appropriateness of half-life being 9 day, the authors added some other evaluations using 3 days and 14 days for half-lives; the results were 0.57 and 0.58 respectively and became slightly worse than employing 9 day half-life.

## V. DISCUSSION

As the proposition for estimation, this method hypotheses that every patient has high fever, but in the actuality, some patients do not show high-fever, but suffer from continuous cough, dyspnea and/or severe general fatigue instead, and become suspicious of the infection and PCR test proves their positivity. However, at the timing when this manuscript is written, LINE data other than high-fever was not open and these symptoms without fever could not be analyzed. A further investigation of the influence for the estimation with other additional symptoms is considered indispensable.

Based on the estimation is around 20 thousand positivity by this paper as of April 1, if the rate of heavily affected patients being around 5%, the number of these patients could be not less than 1,000 patients, which is far more from the presumed number of serious patients which could be assumed from 5,438 patients confirmed test positivity as of April 12. This discrepancy would suggest the tentative consensus of 5% of developing very severe disease would be somewhat exaggerating the actuality.

Meanwhile, in order for our society to achieve so-called “herd immunity”,^11^ in general, 60% to 70% of the lay people should experience the infection. Taking 60% as an example, 75.6 million people nationwide calculated from 126 million population (approximate number by Ministry of Internal Affairs and Communications as of March1, 2020), or 8.4 million Tokyo residents calculated from 14 million residents (approximate number by Tokyo Prefectural Government as of January 1, 2020) should have the individual immunity. This consideration would suggest that, at least at present, the prioritization of acquiring the herd immunity as the public health policy might be obliged to be revisited.

As the limitation to the current method, the regression coefficient is not more than around 0.6; while the existence of correlation and the appropriateness of the method could be proved, it could not give a very precise estimation. Although the authors did not perform the detailed evaluation in terms of the accuracy of the methodology, the estimated value of twenty thousand includes the statistical error of approximately ten thousand. Despite this level of exactitude, this is the first ever paper which successfully determined the true number of the infected nationwide in Japan, the medical/social significance of this evaluation remains huge. A further investigation using the more accumulated statistical data and the continuous revision of this analysis would enhance the accuracy of this estimation method to a large extent, which will serve to determine the national/regional policy against this evolving enemy to the mankind.

## VI. CONCLUSION

The authors estimated the true number of the infected people nationwide in Japan by investigating the correlation over prefectures using the LINE questionnaire and PCR test results. As of April 1, 2020, approximately twenty thousand people were considered infected. As the limitation to the current methodology, the estimated value may include the statistical error of approximately ten thousand. Despite this, medial/social significance of determining the true number of the infected is large. A further investigation with more data and the improvement of methodology would provide with more accurate estimation

## Data Availability

All the data used for this work is available as of today April 15, 2020.

## ACKNOWLEDGMENTS

The authors exhibit sincere appreciation to LINE Corporation^12^ and J.A.G JAPAN Corporation^4^ for their respective disclosure of the precious data. We would also thank Mr. Hiroaki Takata and Dr. Hiroshi Ota for their insights throughout our work.

